# ChatGPT as a bioinformatic partner

**DOI:** 10.1101/2024.08.20.24312291

**Authors:** Gianluca Mondillo, Alessandra Perrotta, Simone Colosimo, Vittoria Frattolillo

**Author notes:** Correspondence: Gianluca Mondillo.

## Abstract

The advanced Large Language Model ChatGPT4o, developed by OpenAI, can be used in the field of bioinformatics to analyze and understand cross-reactive allergic reactions. This study explores the use of ChatGPT4o to support research on allergens, particularly in the cross-reactivity syndrome between cat and pork. Using a hypothetical clinical case of a child with a confirmed allergy to Fel d 2 (cat albumin) and Sus s 1 (pork albumin), the model guided data collection, protein sequence analysis, and three-dimensional structure visualization. Through the use of bioinformatics tools like SDAP 2.0 and BepiPRED, the epitope regions of the allergenic proteins were predicted, confirming their accessibility to immunoglobulin E (IgE) and probability of cross-reactivity. The results show that regions with high epitope probability exhibit high surface accessibility and predominantly coil and helical structures. The construction of a phylogenetic tree further supported the evolutionary relationships among the studied allergens. ChatGPT4o has demonstrated its usefulness in guiding non-specialist researchers through complex bioinformatics processes, making advanced science accessible and improving analytical and innovation capabilities.

## 1. Introduction

ChatGPT4o [1] is an advanced language model developed by OpenAI, based on a deep neural network and the Transformer architecture [2]. This model can generate human-like responses to a wide range of requests in the field of Natural Language Processing (NLP), such as text translation, question answering, and text completion, representing a significant evolution in Artificial Intelligence (AI) and NLP. Besides these areas, ChatGPT4o also has great potential applications in bioinformatics, a field that greatly benefits from advanced computational analysis. The use of bioinformatics techniques in the discovery and analysis of new allergens has allowed for better understanding and management of cross-reactive allergic reactions. An example is the cross-reactivity between shrimp tropomyosin and dust mite tropomyosin, caused by the high sequence homology which can reach 82% [3]. Tools like AlgPRED [4] and BepiPRED [5] accurately predict allergenic epitopes, which are crucial for developing personalized immunotherapeutic treatments. The process begins by identifying an allergenic protein, analyzing sequences and structures through vast databases to predict cross-reactivity. Bioinformatics tools enable the rapid examination of large amounts of data, improving the understanding of cross-reactive reactions and guiding the development of diagnostic tests and targeted treatments. Further experimental studies verify the reactivity of the newly identified allergens. In our project, we use ChatGPT4o as a research support tool, helping us formulate research questions, interpret bioinformatics data, and draft scientific documents, bridging advanced computational skills and practical applications in the field of allergies.

## 2. Materials and Methods

To evaluate the effectiveness of ChatGPT4o as a virtual assistant in our project, we chose to analyze a hypothetical clinical case of cross-reactivity syndrome between cat and pork, known as Pork-Cat Syndrome. This cross-reactive allergic reaction occurs when a person allergic to cats also develops an allergic reaction to pork due to the structural similarity between certain proteins present in cats and pigs, particularly Fel d 2 (cat albumin) and Sus s 1 (pork albumin). People affected by this syndrome usually exhibit allergic symptoms such as hives, swelling, or difficulty breathing after consuming pork [6]. The diagnosis and management of the syndrome require specific allergy tests and, in some cases, consultation with an allergist. We chose a fictitious clinical case of an 8-year-old boy, 25 kg, with a confirmed allergy to cat dander via skin prick test (SPT), positive for the molecular test for Fel d 2, who developed a skin allergic reaction about an hour after eating cooked pork. Subsequent molecular tests revealed an allergy to Sus s 1. This clinical case was selected because it represents a significant example of cross-reactivity between Fel d 2 and Sus s 1, demonstrating how allergies can manifest through cross-reactive reactions between different species. Pork-Cat Syndrome highlights the complexity of allergenic interactions and the importance of bioinformatics tools in their identification and understanding. Furthermore, this case is well studied and documented in scientific literature, allowing us to accurately verify the results obtained. Since we had no prior expertise in bioinformatics, we involved ChatGPT4o to help us better understand the necessary steps and concepts to address. We started by using prompt engineering techniques [7], formulating specific textual inputs (prompts) to obtain the best results from ChatGPT4o. In this case, we used the “Role Playing” technique [8], programming ChatGPT4o to assume the role of a bioinformatics expert with experience in pediatric allergology. Through this technique, the chatbot answered our questions and provided information with the language, tone, and depth of knowledge typical of a professional in the field. This approach allowed us to receive detailed and targeted responses, similar to those we would have obtained by consulting a real expert, thus facilitating our project even without prior specialized training. ChatGPT4o promptly provided us with a detailed list of steps to follow, which we summarize below:

1. **Identify the Key Allergen**: ChatGPT4o indicated that we should focus on the allergens Fel d 2 and Sus s 1.
2. **Data Collection**: ChatGPT4o guided us in collecting the amino acid sequences of the proteins in FASTA format, suggesting the use of databases such as Uniprot [9] and NCBI Protein Database [10]. We chose Uniprot for its user-friendly interface.
3. **Use of Bioinformatics Tools**: To analyze the protein sequences and assess the degree of homology, ChatGPT4o recommended using “SDAP 2.0 - Structural Database of Allergenic Proteins” [11] a bioinformatics database designed to study allergenic proteins. We uploaded the FASTA file of Sus s 1 and performed a homology search.
4. **Visualization of Three-Dimensional Structures**: ChatGPT4o suggested using PyMol [12] to visualize and assess the three-dimensional structures of the allergens, indicating the need to download the PDB (Protein Data Bank) files from Uniprot and perform alignments between the proteins to determine spatial similarity.
5. **Epitope Prediction**: ChatGPT4o indicated BepiPred for predicting B-cell epitopes recognizable by IgE starting from the protein sequences in FASTA format.

Following these steps, guided by ChatGPT4o’s instructions, we were able to explore the complex field of bioinformatics and evaluate the model’s ability to support us in scientific research, even without specialized training in the field.

## 3. Results

Here is what the model instructed us to do in detail.

1. Identify the Key Allergen: ChatGPT4o correctly indicated which allergens to focus our attention on, namely cat albumin (Fel d 2) and pork albumin (Sus s 1).
2. Data Collection: ChatGPT4o advised us to collect the amino acid sequences of Fel d 2 and Sus s 1 in FASTA format, pointing out two useful sites for this purpose: Uniprot and NCBI Protein Database. We chose Uniprot for its intuitive graphical interface, thus obtaining the amino acid sequences of both allergens. The FASTA format is a text file used in bioinformatics to represent sequences of amino acids or nucleotides, where these are illustrated using alphabetic letters to indicate the various amino acid residues or nucleotide bases.
3. Use of Bioinformatics Tools: To analyze the amino acid structures of the allergens, assess the degree of homology, and predict possible cross-reactivity, ChatGPT4o recommended using SDAP 2.0. This database and set of bioinformatics tools is designed to facilitate the study of allergenic proteins, allowing the identification and characterization of the potential allergenicity of proteins based on their structure and sequence.

By uploading the FASTA format of an allergen, SDAP 2.0 allows sequence homology searches using the BLAST algorithm to identify similar sequences in the database. In our case, the chatbot instructed us to submit the FASTA file of either Fel d 2 or Sus s 1. We arbitrarily chose the sequence of Sus s 1 to search the database for molecules with similar amino acid sequences, resulting in a table with the search results (Table 1).

**Table 1.** Alignment between homologous proteins to Sus S 1. Alignment made with FASTA version 36.3.8. As explained in the FASTA manual, the bit score is equivalent to the bit score reported by BLAST. A 1 bit increase in score corresponds to a 2-fold reduction in expectation, and a 10-bit increase implies 1000-fold lower expectation. Sequences with E values < 0.01 are almost always homologous. All FASTA search sequence alignment is printed in Blast format where Query is input sequence, and Sbjct is sequence found in the database. Key: protein name (protein type, scientific name of species, common name). Fel d 2 (Albumin, Felis catus, Domestic cat), Bos d 6 (Albumin, Bos Taurus, Cow), Can f 3 (Albumin, Canis lupus familiaris, Domestic dog), Equ c 3 (Albumin, Equus caballus, Horse), Gal d 5 (Albumin, Gallus gallus domesticus, Chicken), Sus s 1 (Albumin, Sus scrofa, Domestic pig).

| UniProt Name | Name | Lenght | Opt | Bits Score | E-Value | Identities (%) | Positives (%) | Gaps (%) |
|---|---|---|---|---|---|---|---|---|
| P49064 | Fel d 2 | 608 | 3386 | 778.0 | 0 | 483/607 (79%) | 546/607 (89%) | 1/607 (0%) |
| P02769 | Bos d 6 | 607 | 3451 | 792.0 | 0 | 485/607 (79%) | 549/607 (90%) | 0/607 (0%) |
| P49822 | Can f 3 | 608 | 3379 | 776.4 | 0 | 476/606 (78%) | 543/606 (89%) | 1/606 (0%) |
| CAA52194 | Equ c 3 | 607 | 3295 | 757.3 | 6.6e-220 | 464/607 (76%) | 531/607 (87%) | 0/607 (0%) |
| CAA43098 | Gal d 5 | 615 | 2032 | 469.4 | 3.1e-133 | 267/606 (44%) | 408/606 (67%) | 5/606 (0%) |
| P08835 | Sus s 1 | 607 | 4162 | 954.9 | 0 | 607 (100%) | 607 (100%) | 0/607 (0%) |

Not understanding the meaning of the tabulated values, we asked ChatGPT4o for assistance in explaining the significance of these results:

**UniProt Name**: This is the unique identifier code assigned to the protein in the UniProt database.

**Name**: This is the common name by which the protein is known.

**Length**: This refers to the length of the protein, measured by the number of amino acids it contains.

**Opt**: The optimized alignment score, which reflects the overall similarity between the sequences being compared.

**Bits Score**: This score helps evaluate the quality of the alignment. A higher bit score indicates a better alignment.

**E-Value**: The E-value shows how many alignments are expected by chance that would have a score equal to or better than the observed score, given the size of the database and the query. A lower E-value suggests a more statistically significant alignment.

**Identities (%)**: This percentage indicates how many amino acid positions are exactly the same between the two aligned sequences.

**Positives (%)**: This percentage reflects the similarities, including amino acids that have been conservatively substituted and have similar chemical properties.

**Gaps (%)**: This percentage shows where sequences have been extended or truncated to optimize the alignment.

**Data Interpretation**: Proteins such as Fel d 2, Bos d 6, and Can f 3 show high identity and positivity (over 78% and 89%, respectively), which suggests they have a high similarity and possible cross-reactivity. On the other hand, Equ c 3 has good identity and positivity (over 76% and 87%), indicating significant similarity with some differences. In contrast, Gal d 5 shows low identity (44%) and positivity (67%), indicating lesser similarity and lower probability of cross-reactivity.

**Final Considerations**: Proteins with high similarity, such as Fel d 2, Bos d 6, and Can f 3, are good candidates for studies on allergic cross-reactivity. Conversely, Gal d 5, which shows lower similarity, has a lower probability of cross-reactivity but is useful for understanding protein diversity.

We considered all allergens that, according to the BLAST algorithm on SDAP 2.0, are cross-reactive with Sus s 1, including Gal d 5 as a negative control.

### 4. Visualization of Three-Dimensional Structures

We asked how to visualize and evaluate the degree of overlap of the three-dimensional structures of allergens, and ChatGPT4o recommended using PyMol (version 3.0.3). PyMol is a powerful open-source molecular visualization software widely used in structural biology and bioinformatics. This tool allows for the precise and detailed visualization and manipulation of three-dimensional structures of molecules, such as proteins and nucleic acids. It is particularly useful for creating images and animations of molecular structures, facilitating the understanding of interactions between atoms and molecules within a biological system.

PyMol supports various file formats, including the PDB format, which contains atomic coordinates of molecular structures. With PyMol, it is possible to color molecules based on different properties, such as atom type, polypeptide chain, or secondary structure, thereby enhancing visual analysis capabilities. This tool is essential for researchers who wish to examine the three-dimensional structure of allergens and their potential interactions in detail, facilitating the identification of binding sites and epitopes that may be relevant for allergenic cross-reactivity.

To evaluate whether there were overlaps in the three-dimensional structures of the allergens, ChatGPT4o also advised us to use PyMol, which, with the “align protein x, protein y” function, allows for the acquisition of data on the spatial similarity of the analyzed proteins. To perform this evaluation, we needed the PDB files of each of the allergens under study, which are easily downloadable from Uniprot.

We loaded these files into PyMol and began alignment evaluations between the various allergens. For each alignment, PyMol provides a series of data, including the Root Mean Square Deviation (RMSD) [13], which indicates the similarity between the three-dimensional protein structures.

In particular, RMSD is a quantitative measure used to assess the structural similarity between two molecules, especially proteins, after they have been aligned. It is calculated as the square root of the average of the squares of the differences between the corresponding atomic positions in the two structures. In practice, RMSD provides a numerical value that reflects how closely two overlaid molecular structures match each other.

A low RMSD value indicates that the two structures are very similar, while a high value suggests a significant structural difference. RMSD is widely used in structural biology to compare protein models, validate structural predictions, and analyze conformational movements.

The results were put in a table that sorted the protein alignments by RMSD value (Table 2). In Figure 1, the graphical visualization of the overlay of different protein structures performed using PyMol is shown.

**Table 2.** RMSD Values and Number of Aligned Atoms Between Studied Allergenic Proteins. RMSD measures the structural similarity between two proteins, with lower values indicating greater similarity. The numbers in parentheses indicate the number of atoms used in the structural alignment for the RMSD calculation. A higher number of aligned atoms suggests that a larger portion of the proteins was overlapped during the comparison. However, the actual similarity between the structures also depends on the RMSD value: a low RMSD value with a high number of aligned atoms indicates a strong structural similarity, while a high RMSD value may indicate significant structural differences even if many regions are aligned. For example, the comparison between Fel d 2 and Sus s 1 shows an RMSD of 1.036 with 3421 aligned atoms, indicating a moderate difference in their three-dimensional structure. The comparison between Can f 3 and Equ c 3 shows an RMSD of 0.739 with 3307 aligned atoms, indicating greater structural similarity between these two proteins. On the other hand, Fel d 2 and Gal d 5 have an RMSD of 2.084 with 3097 aligned atoms, suggesting significant structural differences between these proteins. These data are fundamental for understanding the evolutionary and structural relationships between allergenic proteins and for identifying potential common epitopes that could cause cross-reactive reactions.

| RMSD<br>(Aligned Atoms) | Fel d 2 | Sus s 1 | Can f 3 | Equ c 3 | Gal d 5 | Bos d 6 |
|---|---|---|---|---|---|---|
| Fel d 2 | 0 (4391) | 1.036 (3421) | 0.749 (3270) | 0.957 (3405) | 2.084 (3097) | 1.060 (3659) |
| Sus s 1 | 1.036 (3421) | 0 (9725) | 0.723 (3854) | 0.998 (3408) | 2.055 (5976) | 0.863 (3500) |
| Can f 3 | 0.749 (3270) | 0.723 (3854) | 0 (4807) | 0.739 (3307) | 1.915 (3282) | 0.930 (3580) |
| Equ c 3 | 0.957 (3405) | 0.998 (3408) | 0.739 (3307) | 0 (4510) | 1.884 (3005) | 0.941 (3572) |
| Gal d 5 | 2.084 (3097) | 2.055 (5976) | 1.915 (3282) | 1.884 (3005) | 0 (9646) | 1.963 (3112) |
| Bos d 6 | 1.060 (3659) | 0.863 (3500) | 0.930 (3580) | 0.941 (3572) | 1.963 (3112) | 0 (9340) |

**Figure 1.**
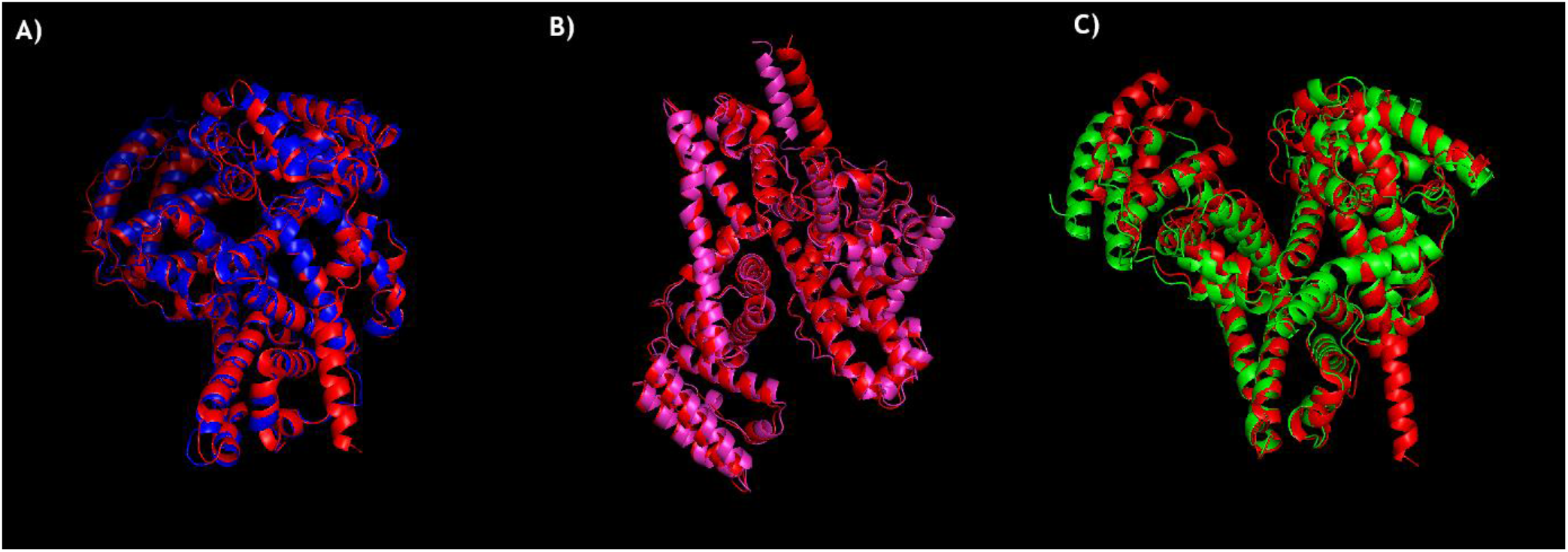
Alignment of Different Three-Dimensional Protein Structures in PyMol: A) Fel d 2 (in blue) vs Sus s 1 (in red), B) Sus s 1 (in red) vs Can f 3 (in pink), C) Sus s 1 (in red) vs Gal d 5 (in green)

### 5. Epitope Prediction

We asked ChatGPT4o if it was possible to predict, based on the protein structures of the allergens under study, which epitopes could be recognized by IgE antibodies. ChatGPT4o recommended using BepiPRED, a machine learning algorithm specifically adapted for predicting epitopes recognizable by antibodies from protein sequences. This tool helps identify critical regions based on the amino acid sequence of the protein, providing crucial information for immunological research, vaccine development, and allergenicity studies. We uploaded the FASTA sequences of all our allergens into BepiPred, from which we obtained data that were subsequently analyzed with the help of ChatGPT4o.

The data analysis provided the following results:

1. Relative Surface Accessibility: The average surface accessibility for regions with high epitope probability is 0.416, with a minimum of 0.025 and a maximum of 0.813. This indicates that regions with high epitope probability tend to have good surface accessibility, making them potentially recognizable by antibodies.
2. Helix Probability: The average probability of helical structures is 0.421, suggesting a significant presence of helical structures in the epitope regions. The variability is high, with a maximum of 0.988.
3. Sheet Probability: The average probability of β-sheet structures is very low, at 0.018, indicating that these structures are less common in regions with high epitope probability.
4. Coil Probability: The average probability of coil structures is 0.562, suggesting that these structures are predominant in the epitope regions.
5. Exposed Percentage: 74.49% of residues with high epitope probability are classified as exposed, while 25.51% are buried. This is a strong indicator that most of these regions are accessible to the immune system, essential for antibody recognition.

Figures 2 and 3 display graphs representing the epitope probability for each analyzed protein. These graphs were generated using the Python (version 3.12.3) libraries Pandas (version 2.0.3) and Matplotlib (version 3.7.2).

**Figure 2.**
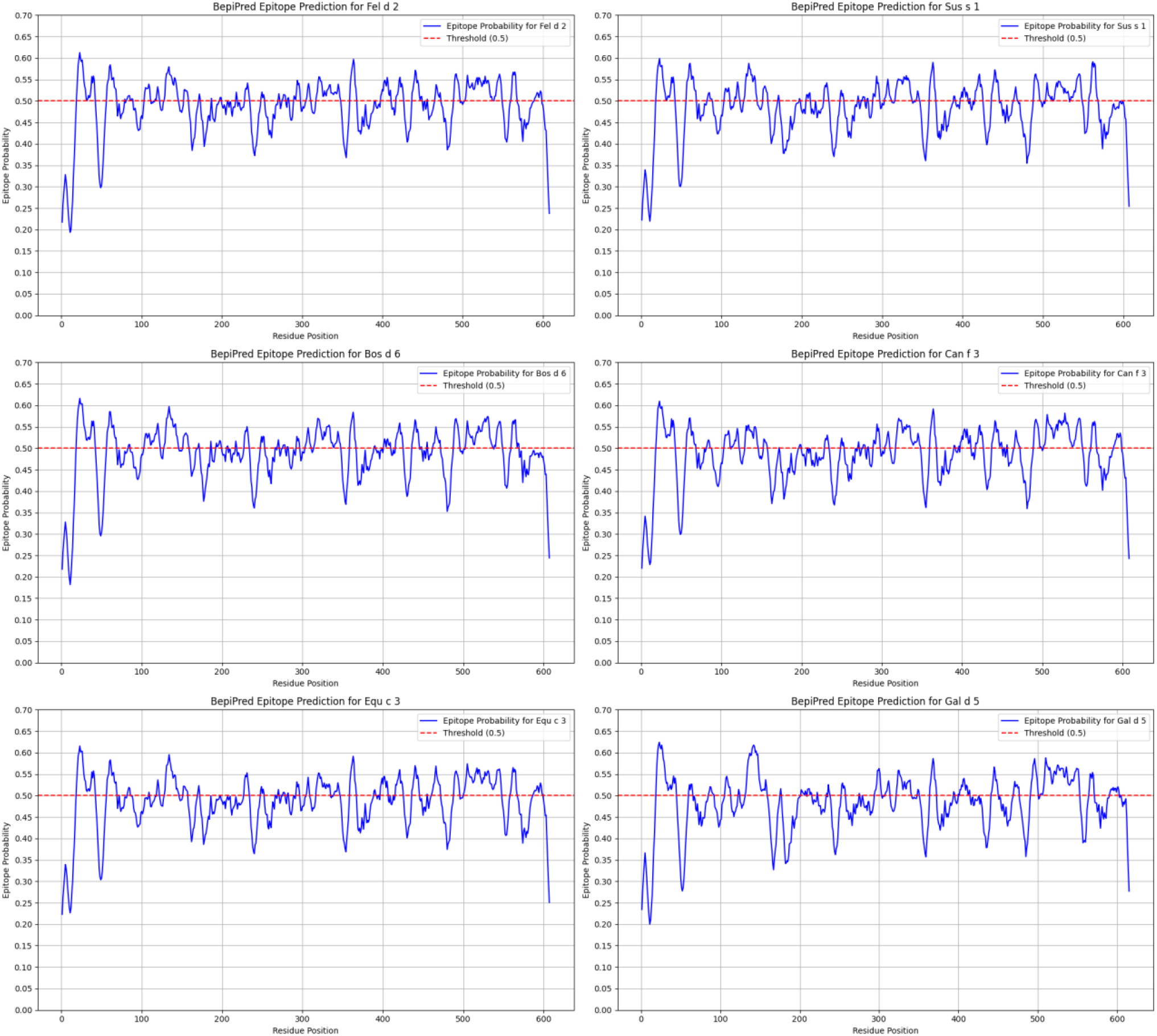
B-cell Epitope Prediction along the Protein Sequence of Fel d 2 (cat), Sus s 1 (pig), Bos d 6 (cow), Can f 3 (dog), Equ c 3 (horse), Gal d 5 (chicken), using BepiPRED. The blue line represents the epitope probability for each amino acid residue, while the dashed red line indicates the threshold of 0.5, a value suggested by ChatGPT4o. Regions above the threshold are considered potential epitopes.

**Figure 3:**
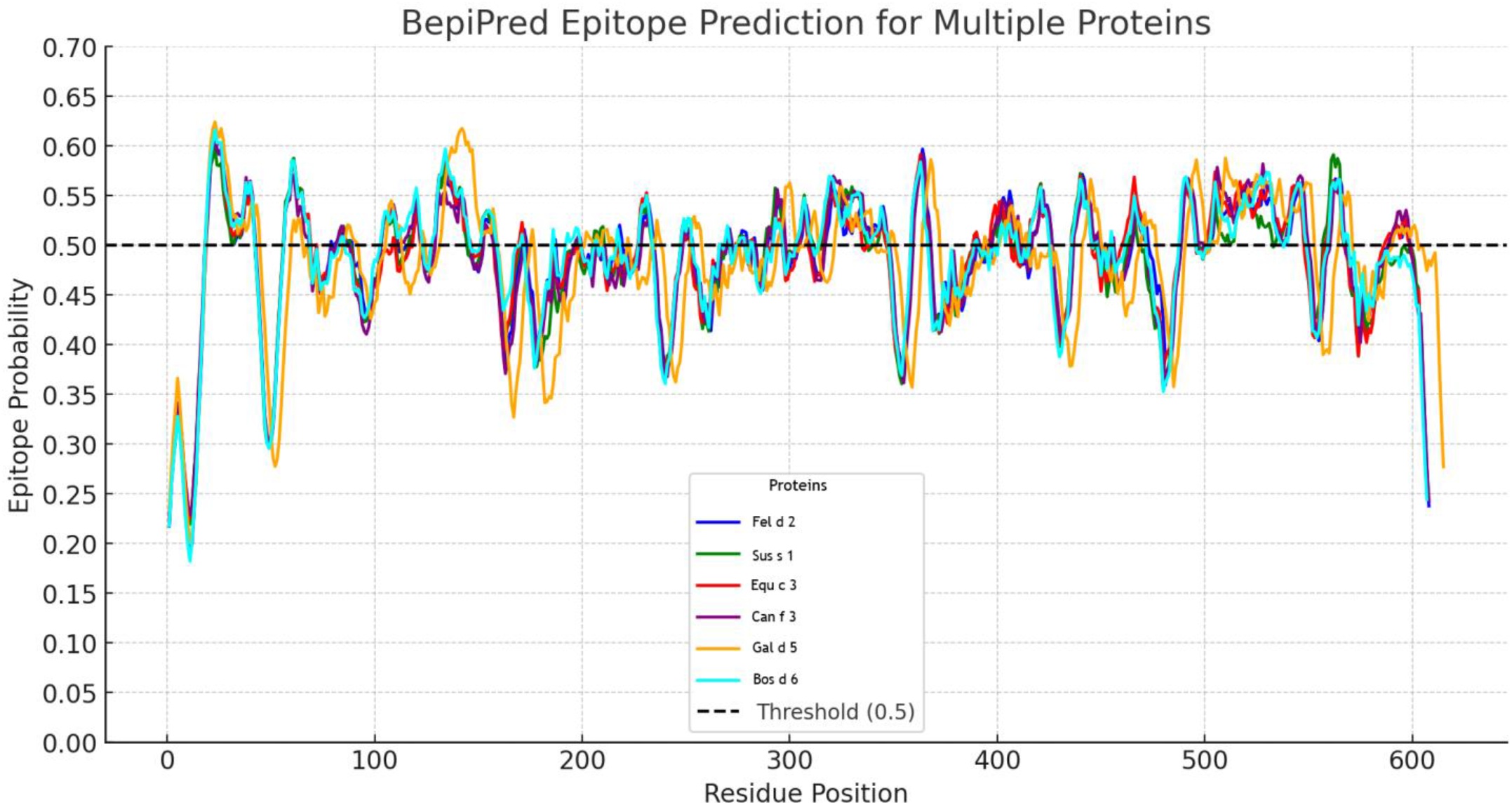
B-cell Epitope Prediction along the Protein Sequences of Various Allergenic Proteins using BepiPRED. The analyzed proteins include Fel d 2 (cat, blue), Sus s 1 (pig, green), Equ c 3 (horse, red), Can f 3 (dog, purple), Gal d 5 (chicken, orange), and Bos d 6 (cow, cyan). The dashed black line represents the threshold of 0.5 for epitope probability. Regions of the protein sequence that exceed this threshold are considered potential epitopes. Overlaps between the colored lines suggest potential cross-reactivity among these allergenic proteins, while fluctuations along the sequences indicate variations in epitope probability.

These results confirm that regions with high B-cell epitope probability show high surface accessibility and a predominance of coil structures, with a significant presence of helical structures. The vast majority of these residues are exposed, making them ideal candidates for interactions with specific antibodies. This is a crucial aspect for immune recognition and can have important implications for vaccine design or allergenicity studies.

Lastly, we asked how to generate a phylogenetic tree of the proteins under analysis, and the model recommended the website “phylogeny.fr” [14] which, by uploading the protein sequences in FASTA format, generates a phylogenetic tree, as shown in Figure 4.

**Figure 4:**
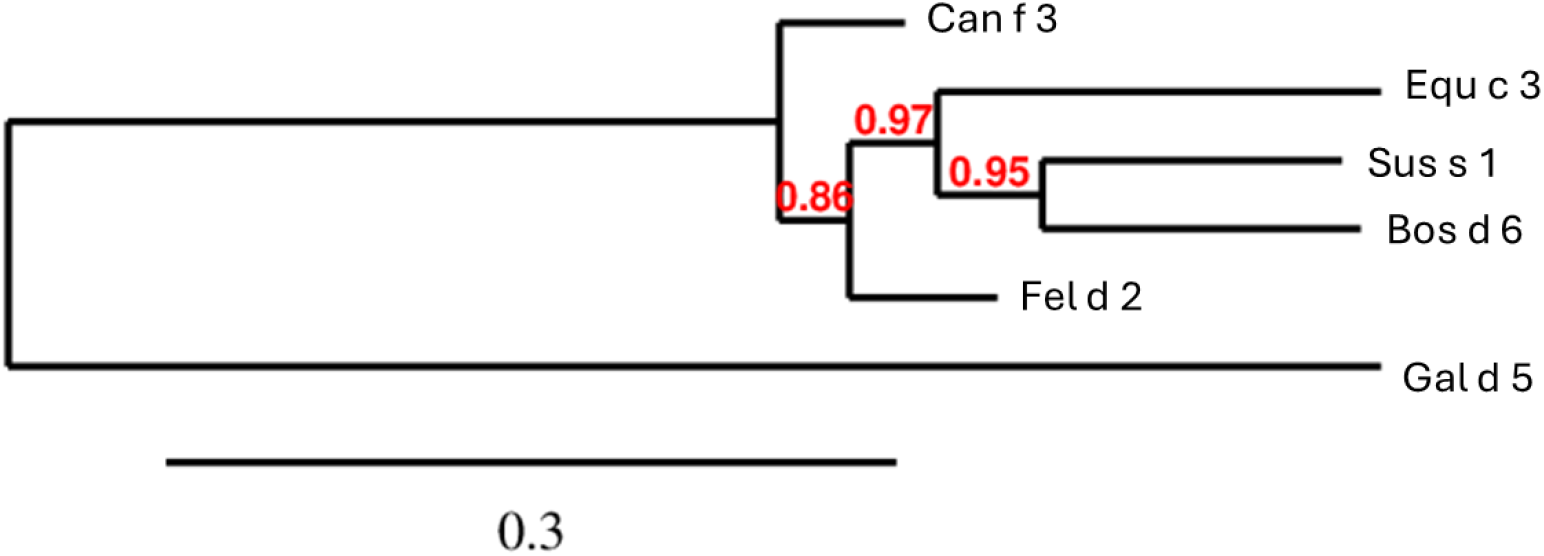
Phylogenetic Tree of the Analyzed Allergenic Proteins Constructed Using the Neighbor-Joining Method. The analyzed species include Fel d 2 (cat), Can f 3 (dog), Bos d 6 (cow), Equ c 3 (horse), Sus s 1 (pig), and Gal d 5 (chicken). The numerical values in red represent the bootstrap values, which indicate the robustness of the nodes in the phylogenetic tree. A high bootstrap value suggests greater reliability of the represented evolutionary relationship. The scale bar at the bottom with a value of “0.3” represents evolutionary distance: a branch length corresponding to this scale indicates a 30% difference between the protein sequences. The tree shows a close relationship between Fel d 2, Can f 3, Equ c 3, Sus s 1, and Bos d 6, while Gal d 5 appears to be more evolutionarily distant.

## 4. Discussion

Bioinformatics is making significant advances in understanding and managing cross-reactive allergic reactions. Tools like SDAP 2.0 allow precise identification and characterization of protein allergenicity based on structure and sequence. This is crucial in complex diseases such as cross-reactivity syndrome between Fel d 2 and Sus s 1, where predicting cross-reactive allergic reactions can guide diagnosis and therapy. Studies have shown that proteins with high sequence homology (85% and 71%) exhibit cross-reactivity, whereas those with lower homology (51% and 60%) do not [15]. A homology threshold of 70-80% is indicative of cross-reactivity [16]. Structural similarity analysis using RMSD supports these predictions, with low RMSD values indicating strong structural similarity. RMSD, along with the number of aligned atoms, is crucial for understanding structural similarity and the likelihood of cross-reactivity. In our study, Gal d 5 was used as a negative control to confirm the bioinformatic distance from the other analyzed allergens. The results showed that Gal d 5 does not exhibit significant cross-reactivity, confirming the reliability of our bioinformatics approach. The use of PyMol revealed that proteins with high sequence homology have low RMSD values, confirming structural similarity and the identification of potential cross-reactive allergens. These results improve diagnostic and therapeutic strategies for cross-reactive allergies, highlighting the importance of bioinformatics techniques. The initial approach often starts with bioinformatics to filter and identify sequences with high homology, followed by experimental studies to validate predictions. Tools like ChatGPT4o are invaluable, especially for those without specialized training in bioinformatics [17]. ChatGPT4o helps analyze and synthesize information, guiding users through complex bioinformatics processes. Its ability to act as an intermediary between advanced computational skills and practical applications makes advanced science accessible even to non-specialists, revolutionizing scientific research and the management of complex medical problems [18]. ChatGPT has been used to simplify genomic sequence analysis, phylogenetic tree construction, and adaptation of computer vision algorithms, demonstrating its versatility in supporting researchers without in-depth specialized training [19]. Additionally, ChatGPT has facilitated learning and teaching in bioinformatics, helping students and researchers explore new educational possibilities and develop critical skills [20]. Its utility extends beyond simple search functions, acting as a catalyst for innovation and efficiency, allowing scientists to focus on more creative and analytical aspects of research.

## 5. Conclusions

The integration of AI tools like ChatGPT in the field of bioinformatics offers a significant advantage for research, especially for groups with limited resources or without specialized training. Thanks to ChatGPT’s ability to provide quick and understandable answers on complex topics, even researchers without advanced bioinformatics skills can obtain valuable information for their studies. This tool facilitates access to bioinformatics, allowing for the formulation of research questions, data interpretation, and drafting of scientific documents with greater efficiency. However, for more complex tasks and the interpretation of sophisticated data, the intervention of bioinformatics experts remains indispensable. In summary, the use of ChatGPT and, more generally, AI, represents an important step forward towards the democratization of science, making advanced research more accessible and improving analytical and innovation capabilities even in contexts with limited resources.

## Data Availability

All data produced in the present work are contained in the manuscript

## Author Contributions

all authors have contributed equally.

## Funding

“This research received no external funding”.

## Conflicts of Interest

“The authors declare no conflicts of interest.”

## Notes

### Competing Interest Statement

The authors have declared no competing interest.

### Funding Statement

This study did not receive any funding

## References

1. OpenAI. Hello GPT-4o. Available online: https://openai.com/index/hello-gpt-4o/

2. Vaswani, A.; Shazeer, N.M.; Parmar, N.; Uszkoreit, J.; Jones, L.; Gomez, A.N.; Kaiser, L.; Polosukhin, I. Attention is All you Need. Neural Information Processing Systems 2017.

3. Saetang, J.; Tipmanee, V.; Benjakul, S. In Silico Prediction of Cross-Reactive Epitopes of Tropomyosin from Shrimp and Other Arthropods Involved in Allergy. Molecules 2022, 27, 2667. 10.3390/molecules27092667

4. Sharma, N.; Patiyal, S.; Dhall, A.; Pande, A.; Arora, C.; Raghava, G.P.S. AlgPred 2.0: an improved method for predicting allergenic proteins and mapping of IgE epitopes. Brief Bioinform. 2021, 22. doi: 10.1093/bib/bbaa294. PMID: 33201237.

5. Clifford, J.N.; Høie, M.H.; Deleuran, S.; Peters, B.; Nielsen, M.; Marcatili, P. BepiPred-3.0: Improved B-cell epitope prediction using protein language models. Protein Sci. 2022, 31. doi: 10.1002/pro.4497. PMID: 36366745; PMCID: PMC9679979.

6. Drouet, M.; Boutet, S.; Lauret, M.G.; Chène, J.; Bonneau, J.C.; Le Sellin, J.; Hassoun, S.; Gay, G.; Sabbah, A. Le syndrome porc-chat ou l’allergie croisée entre viande de porc et épithélia de chat (1re partie) [The pork-cat syndrome or crossed allergy between pork meat and cat epithelia (1)]. Allerg Immunol (Paris). 1994, 26, 166-8, 171-2. PMID: 8086105.

7. IBM. What is prompt engineering? Available online: https://www.ibm.com/topics/prompt-engineering

8. Shanahan, M.; McDonell, K.; Reynolds, L. Role play with large language models. Nature 2023, 623, 493–498. doi: 10.1038/s41586-023-06647-8. PMID: 37938776.

9. UniProt. Available online: https://www.uniprot.org/

10. National Center for Biotechnology Information (NCBI) [Internet]. Bethesda (MD): National Library of Medicine (US), National Center for Biotechnology Information; [1988] – [cited 2024 Jul 15]. Available online: https://www.ncbi.nlm.nih.gov/

11. Schein, C.H.; Negi, S.S.; Braun, W. Still SDAPing Along: 20 Years of the Structural Database of Allergenic Proteins. Front. Allergy 2022, 3, 863172. doi: 10.3389/falgy.2022.863172.

12. Schrödinger, LLC. PyMOL The PyMOL Molecular Graphics System, Version 3.0. Available online: https://pymol.org/

13. Kufareva, I.; Abagyan, R. Methods of protein structure comparison. Methods Mol Biol. 2012, 857, 231–57. doi: 10.1007/978-1-61779-588-6_10. PMID: 22323224; PMCID: PMC4321859.

14. Robust Phylogenetic Analysis For The Non-Specialist. Available online: https://www.phylogeny.fr/

15. Westwood, G.S.; Huang, S.W.; Keyhani, N.O. Molecular and immunological characterization of allergens from the entomopathogenic fungus Beauveria bassiana. Clin Mol Allergy 2006, 4, 12. doi: 10.1186/1476-7961-4-12.

16. Cantillo, J.F.; Puerta, L.; Fernandez-Caldas, E.; et al. Tropomyosins in mosquito and house dust mite cross-react at the humoral and cellular level. Clin Exp Allergy 2018, 48, 1354–1363.

17. Shue, E.; Liu, L.; Li, B.; Feng, Z.; Li, X.; Hu, G. Empowering Beginners in Bioinformatics with ChatGPT. bioRxiv [Preprint]. 2023, doi: 10.1101/2023.03.07.531414.

18. Wang, J.; Cheng, Z.; Yao, Q.; Liu, L.; Xu, D.; Hu, G. Bioinformatics and biomedical informatics with ChatGPT: Year one review. ArXiv [Preprint]. 2024. PMID: 38562449; PMCID: PMC10984005.

19. Aden, D.; Zaheer, S.; Khan, S. Possible benefits, challenges, pitfalls, and future perspective of using ChatGPT in pathology. Rev Esp Patol. 2024, 57, 198–210. doi: 10.1016/j.patol.2024.04.003.

20. Magalhães Araujo, S.; Cruz-Correia, R. Incorporating ChatGPT in Medical Informatics Education: Mixed Methods Study on Student Perceptions and Experiential Integration Proposals. JMIR Med Educ. 2024, 10. doi: 10.2196/51151. PMID: 38506920; PMCID: PMC10993110.

